# Studies on the level of neutralizing antibodies produced by inactivated COVID-19 vaccines in the real world

**DOI:** 10.1101/2021.08.18.21262214

**Authors:** Haiying Zhang, Yuyuan Jia, Ying Ji, Xu Cong, Yan Liu, Ruifeng Yang, Xiangsha Kong, Yijun Shi, Ling Zhu, Zhenyu Wang, Wei Wang, Ran Fei, Feng Liu, Fengmin Lu, Hongsong Chen, Huiying Rao

## Abstract

**Background:** Although effective vaccines have been developed against COVID-19, the level of neutralizing antibodies (Nabs) induced after vaccination in the real world is still unknown. To evaluate the level and persistence of NAbs induced by two inactivated COVID-19 vaccines in China.

**Methods and findings:** Serum samples were collected from 1,335 people aged 18 and over who were vaccinated with COVID-19 inactivated vaccine in Peking University People’s Hospital from January 19 to June 23, 2021, for detection of COVID-19 antibodies. The WHO standard of SARS-CoV-2 NAbs was detected. The coefficients of variation between the detection results and the true values of the NAbs detected by the WHO standard were all lower than the WHO international standard 3% after the dilution of the original and the dilution of the theoretical concentrations of 500 IU/mL, 250 IU/mL, 125 IU/mL, 72.5 IU/mL, 36.25 IU/mL and 18.125 IU/mL. On day 11-70, the positive rate of NAbs against COVID-19 was 82% to 100%; From day 71 to 332, the positive rate of NAbs decreased to 27%. The level of NAbs was significantly higher at 3-8 Weeks than at 0-3 Weeks. There was a high linear correlation between NAbs and IgG antibodies in 1335 vaccinated patients. NAbs levels were decreased in 31 of 38 people (81.6%) at two time points after the second dose of vaccine. There was no significant difference in age between the group with increased and decreased neutralizing antibody levels (*χ*2 =-0.034, *P*>0.05). The positive rate of NAbs in the two-dose vaccine group (77.3%) was significantly higher than that in the one-dose group (18.1%), with statistical difference (*χ*2=312.590, *P*<0.001). A total of 206 people who were 11-70 days after receiving the second dose were tested and divided into three groups: 18-40 years old, 41-60 years old and >60 years old. The positive rates of NAbs in three groups (18-40 years old, 41-60 years old and >60 years old) were 95.14%, 78.43% and 81.8%, respectively. The positive rate of NAbs was significantly higher in 18-40 years old than in 41-60 years old (*χ*2=12.547, *P* <0.01). The titer of NAbs in 18-40 years old group was significantly higher than that in 41-60 years old group (t=-0.222, *P* <0.01). The positive rate of NAbs in male group (89.32%) was lower than in female (91.26%), but there was no significant difference (*χ*2=0.222, *P* >0.05).

**Conclusions:** The positive rate of NAbs was the highest from 10 to 70 days after the second dose of vaccine, and the positive rate gradually decreased as time went by. There was a high linear correlation between COVID-19 NAbs and IgM/IgG antibodies in vaccinators, suggesting that in cases where NAbs cannot be detected, IgM/IgG antibodies can be detected instead. The level of NAbs produced after vaccination was affected by age, but not by gender. The highest levels of NAbs were produced between shots 21 to 56 days apart, suggesting that 21 to 56 days between shots is suitable for vaccination.

**Author summary:** *Why was this study done?:*

1. At present, the inactivated vaccines that have been approved to market in China have passed clinical trials to prove their effectiveness and safety. But the level of neutralizing antibodies induced by vaccination in the real world remains unclear.
2. Serological testing for neutralizing antibodies against COVID-19 is important for assessing vaccine and treatment responses and comparing multiple drug candidates. We assessed the levels of neutralizing antibodies produced in populations receiving inactivated vaccines and assessed the persistence of these vaccines in producing COVID-19 neutralizing antibodies in healthy adults.

*What did the researchers do and find?:*

1. We collected serum samples from 1,335 people aged 18 and above who had received COVID-19 vaccine in Peking University People’s Hospital, and divided them into two groups according to one dose of inactivated vaccine and two doses of inactivated vaccine.
2. Our study found that the positive rate of NAbs was 66.2% in adults who received one or two doses of inactivated vaccine and 77.3% in adults who received two doses of inactivated vaccine in the real world.
3. From 11 to 70 days after the second dose of vaccine, the positive rate of neutralizing antibodies against COVID-19 was 82-100%; On days 71-332, the positive rate of neutralizing antibodies decreased to 27%.
4. The titer and the positive rate of NAbs in 18-40 years old group were significantly higher than that in 41-60 years old group.

*What do these findings mean?:*

1. What is novel is we observed that in the real world, the positive rate of neutralization antibody was the highest at 10 to 70 days after the second vaccination, and with the extension of the vaccination time, the positive rate of antibody gradually decreased. Therefore, we recommend that the third dose of vaccine be administered at day 61 to day 70 for COVID-19 neutralizing antibodies levels.
2. We observed that there was a high linear correlation between COVID-19 neutralization antibodies and COVID-19 IgM/IgG antibodies in vaccinators, suggesting that in cases where NAbs cannot be detected, COVID-19 IgM/IgG antibodies can be detected instead.
3. In our manuscript, we found that the titer and positive rate of neutralizing antibodies in 18-40 years old group were higher than those in 41-60 years old group. The level of neutralizing antibodies produced after vaccination was affected by age, but not by gender.
4. We also observed that the highest levels of NAbs were produced between shots 21 to 35 days apart, suggesting that 21 to 35 days between shots is suitable for vaccination.

## Introduction

Novel Coronavirus 2019 (SARS-CoV-2) is the causative agent of Novel Coronavirus Pneumonia (COVID-19), which is the cause of the global pandemic. A safe, effective and rapidly deployable vaccine is an effective global measure to control the spread of outbreaks. As virus mutations have been reported recently, an effective vaccine against COVID-19 is urgently needed to control the global COVID-19 pandemic. Currently, more than 280 vaccine candidates are in development worldwide, of which 23 are in phase 3 clinical trials [1]. Inactivated vaccines, such as influenza vaccine, hepatitis B vaccine, poliomyelitis vaccine and DPT vaccine, have been widely studied and widely used to prevent respiratory tract infection due to their good safety [2]. At present, the inactivated vaccines that have been approved for marketing in China have been proved to be effective and safe through clinical trials [3,4,5]. However, after the marketing of vaccines, the clinical protective effect in the case of large-scale vaccination should be continued to be observed, and the protective persistence of vaccines should be studied. The phase 3 clinical trial of inactivated vaccine in China evaluated the efficacy and persistence of inoculation by detecting neutralizing antibody titers [5]. Currently, China has launched inactivated COVID-19 vaccines from Beijing Institute of Biological Products (SINophin), Wuhan Institute of Biological Products (Sinophin) and Sinovac. According to the official website of the National Health Commission, as of August 1, 2021, a total of 16,69,527,000 doses of Novel Coronavirus vaccine have been reported in 31 provinces (autonomous regions and municipalities right under the central government) and Xinjiang Production and Construction Corps [6].

However, the level of production of COVID-19 neutralizing antibodies (NAbs) from inactivated vaccines in the real world has not been confirmed and needs to be evaluated in large samples. Serological testing for NAbs against COVID-19 is important for assessing vaccine and treatment responses and comparing multiple drug candidates. In addition, reliable serological testing is needed to understand the true impact of COVID-19 through sero-epidemiological studies, as most cases are asymptomatic, and mild cases go largely undetected [7]. Virus neutralization tests (NTs) are the gold standard for the detection of NAbs, but they are complex and require BSL3 facilities. In contrast, alternative fully automated chemiluminescence instruments offers the possibility of high-throughput testing in the laboratory. Therefore, we evaluated the crowd of inactivated vaccine inoculation produce NAbs’ level, evaluate the vaccine in healthy adults to generate new NAbs persistence, and analyze Nabs’ level with the patient age, gender, and the relationship between the new crown antibody levels, for vaccine research and development, drug treatment, epidemiology and immune surveillance provide important reference basis.

## Methods

### Study population

Serum samples were collected from 1335 patients aged 18 and over who received one dose of inactivated vaccine or two doses of inactivated vaccine in Peking University People’s Hospital from January 19 to June 23, 2021. They were divided into two groups, 244 patients received one dose of inactivated vaccine and 1,091 patients received two doses of inactivated vaccine. All patients had no history of COVID-19 infection. Two inactivated vaccines were isolated from two patients with SARS-CoV-2 (WIV04 and HB02) in Jinyintan Hospital, Wuhan, China, and used to develop two vaccines (hereinafter referred to as WIV04 and HB02 vaccines), respectively [5]. This study was approved by the ethics committee of the Peking University People’s Hospital.

### Measurement of SARS-COV-2 antibody detection

The indirect method was used for detection. Magnetic particles were coated with 2019-nCoV antigen, and anti-human IgG antibody was labeled with horseradsh peroxidase to prepare enzyme conjugals. The complex catalyzed luminescence substrate to give off photoperites, and the luminescence intensity was proportional to the content of COVID-19 IgG antibody. The serum sample size required was 80μl. When S/CO≥1.00, the result was positive; When S/CO < 1.00, the result was negative. Autolumo A2000 PLUS automatic chemiluminescence immunoanalyzer from Zhengzhou Antu Biological Engineering Co., LTD was used as the supporting instrument for the above reagents.

### Measurement of SARS-CoV-2 neutralizing antibodies detection

Detection of SARS-CoV-2 NAbs: The detection was performed using the principle of competition method, using the principle that the specific binding of ACE2 and RBD proteins would be blocked by the Novel Coronavirus NAbs. The serum sample size required was 80 μl. When the concentration value of sample is ≥30.00 AU/mL, the result is judged as positive, and when the concentration value is < 30.00 AU/mL, the result is judged as negative. Autolumo A2000 PLUS automatic chemiluminescence immunoanalyzer from Zhengzhou Antu Biological Engineering Co., LTD was used as the supporting instrument for the above reagents.

### WHO standard for DETECTION of SARS-COV-2 NAbs

World Health Organization (WHO) first generation international standard for antibodies against COVID-19 (NIBSC Code: 20/136), the international unit concentration (IU/mL) of NAbs was determined, the reference was lyophilized powder, 0.25 mL per tube of standard substance, and the original concentration was 1000IU/mL. The theoretical dilution concentrations were 500 IU/mL, 250 IU/mL, 125 IU/mL, 72.5 IU/mL, 36.25 IU/mL and 18.125 IU/mL. Each sample was tested in parallel for 5 times, and the mean was taken.

### Quality control

Each test batch of the above two kits includes negative quality control products and positive quality control products for COVID-19 antibody/nutralization antibody.

### Statistical analysis

SPSS 21.0 was used for software analysis. The age of patients was measured and normally distributed, denoted by X ± S. *χ*2 test was used for statistical analysis of the count data between the two groups. Statistical analysis of measurement data between the two groups was performed by T test, and *P*<0.050 was considered statistically significant.

## Results

### Analysis of clinical data of 1335 patients

Among the 1335 vaccinated people, there were 644 males and 691 females aged 36.27±12.39 years, and the sex ratio (male/female) was 0.93:1. Among them, there were 243 cases in one-dose vaccine group (age 35.26±11.87 years old), including 120 males and 123 females, with sex ratio (male/female) of 0.98:1; There were 1092 patients (36.50±12.49 years old) in the two-dose vaccine group, including 524 males and 568 females, with a sex ratio of 0.92:1. 570 cases received 21 days between shots, with a median of 21 days between shots (0-77 days); The median number of days between the COVID-19 antibody test date and the second vaccine dose was 50 days (0-332 days)(Table 1).

**Table 1.**
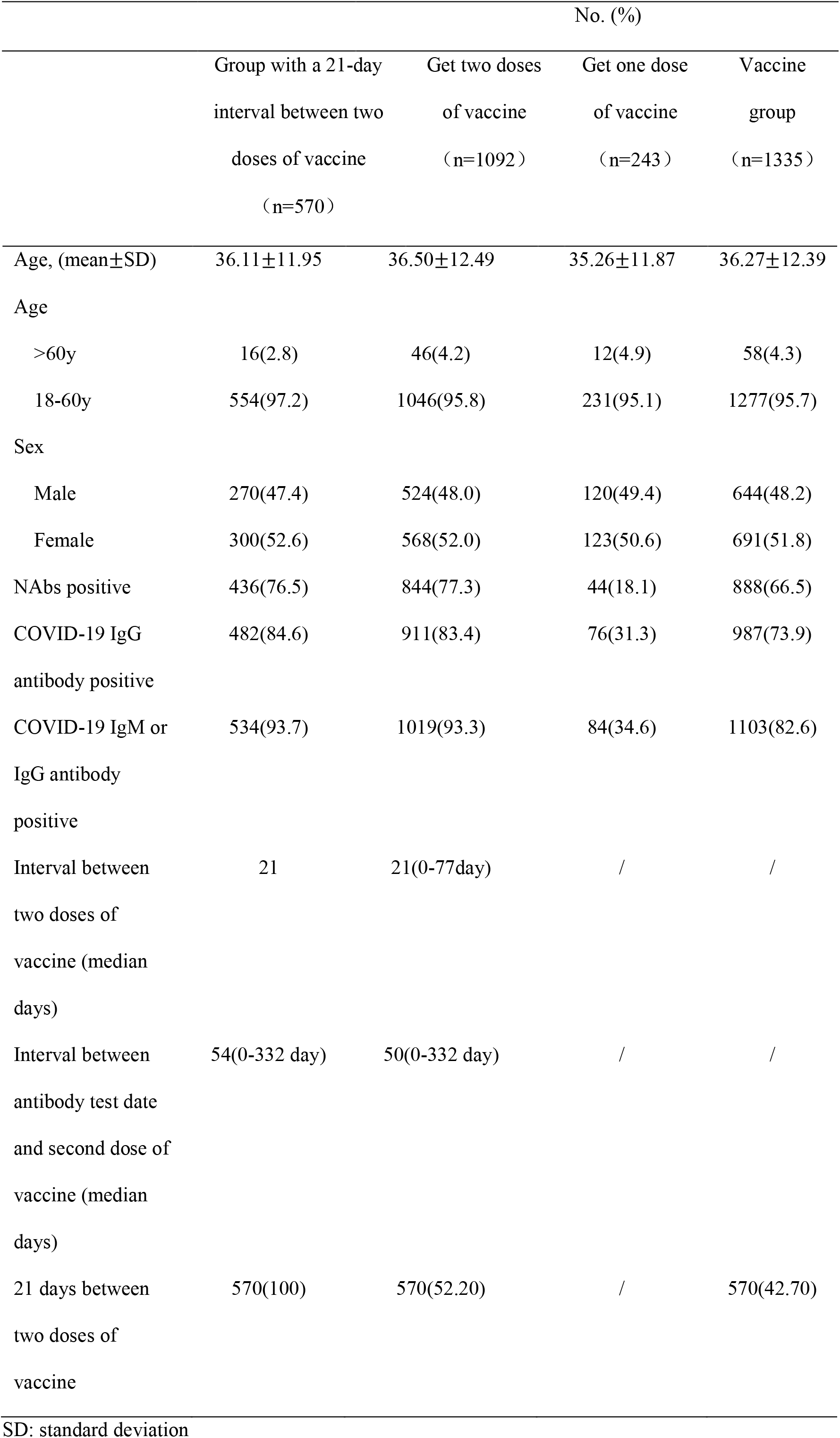
Clinical characteristics of adults vaccinated with the two inactivated vaccines.

|  | Group with a 21-day<br>interval between two<br>doses of vaccine<br>(n=570) | Get two doses<br>of vaccine<br>(n=1092) | Get one dose<br>of vaccine<br>(n=243) | Vaccine<br>group<br>(n=1335) |
| --- | --- | --- | --- | --- |
| Age, (mean±SD) | 36.11±11.95 | 36.50±12.49 | 35.26±11.87 | 36.27±12.39 |
| Age |  |  |  |  |
| >60y | 16(2.8) | 46(4.2) | 12(4.9) | 58(4.3) |
| 18-60y | 554(97.2) | 1046(95.8) | 231(95.1) | 1277(95.7) |
| Sex |  |  |  |  |
| Male | 270(47.4) | 524(48.0) | 120(49.4) | 644(48.2) |
| Female | 300(52.6) | 568(52.0) | 123(50.6) | 691(51.8) |
| NAbs positive | 436(76.5) | 844(77.3) | 44(18.1) | 888(66.5) |
| COVID-19 IgG<br>antibody positive | 482(84.6) | 911(83.4) | 76(31.3) | 987(73.9) |
| COVID-19 IgM or<br>IgG antibody<br>positive | 534(93.7) | 1019(93.3) | 84(34.6) | 1103(82.6) |
| Interval between<br>two doses of<br>vaccine (median<br>days) | 21 | 21(0-77day) | / | / |
| Interval between<br>antibody test date<br>and second dose of<br>vaccine (median<br>days) | 54(0-332 day) | 50(0-332 day) | / | / |
| 21 days between<br>two doses of<br>vaccine | 570(100) | 570(52.20) | / | 570(42.70) |

SD: standard deviation

### WHO standard detection of SARS-COV-2 neutralizing antibody

The WHO standard of SARS-COV-2 neutralizing antibody was detected, and the original concentration of WHO standard was 1000IU/mL. The mean value of our detection results was 1035AU/mL, and the coefficient of variation was 1.3%. The coefficient of variation between the test results and the true values of NAbs of the WHO standard at theoretical concentrations of 500 IU/mL, 250 IU/mL, 125 IU/mL, 72.5 IU/mL, 36.25 IU/mL and 18.125 IU/mL were all lower than the WHO international standard of 3%.

### Changes in neutralization antibody levels at different times after the second dose of inactivated vaccine in the real world

In the 1092 patients in the two-dose inactivated vaccine group, we observed that from day 0 to day 332 after the second dose, with days 11-70, The positive rate of COVID-19 NAbs was 82-100%; during days 71-332, the positive rate of NAbs decreased to 27%. It is recommended to test for neutralizing antibody levels 10-70 days after the second dose(Fig 1-2).

**Fig 1.**
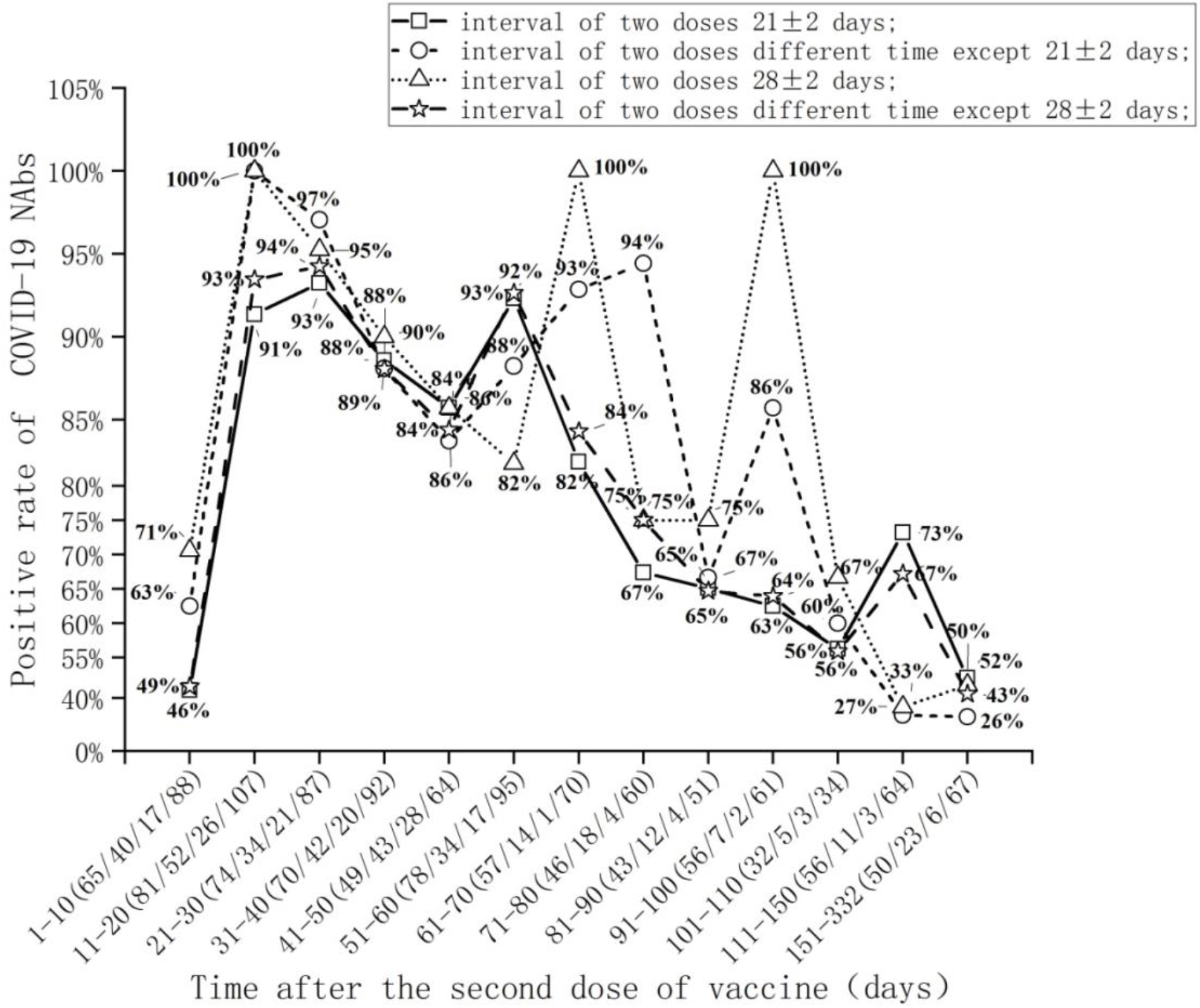
Changes in neutralizing antibodies levels at different times after second dose of inactivated vaccine in the real world

**Fig 2.**
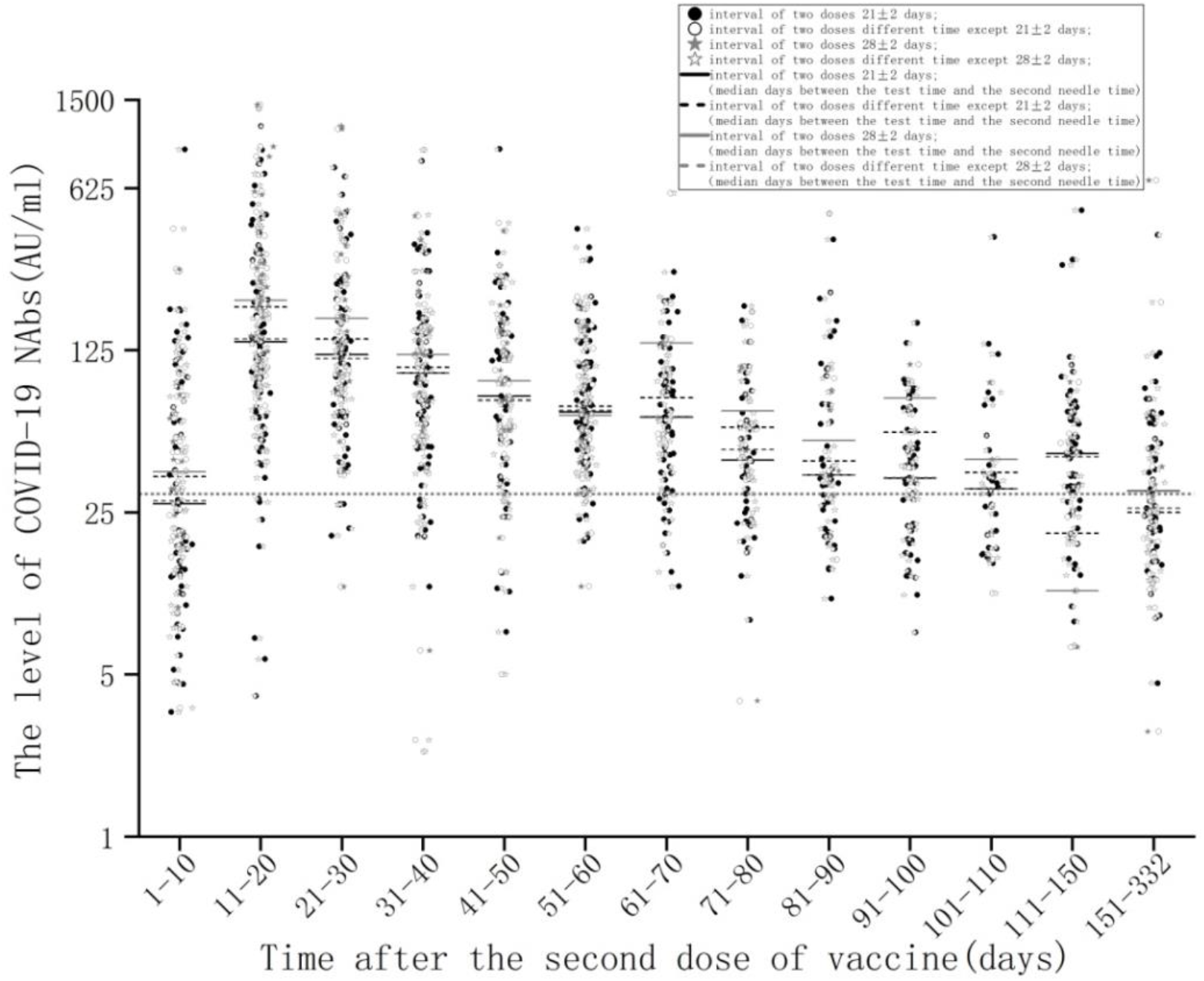
Changes in neutralizing antibodies levels at different times after second dose of inactivated vaccine in the real world. Dotted line indicates cut off value =30 AU/mL;

### COVID-19 antibody levels generated at different time intervals between two doses of vaccine

The interval between two doses of vaccine was from 0 days to 77 days, and the positive rates of NAbs generated at 3-8 weeks intervals between two doses of vaccine were very high (80.8%). The positive rates of COVID-19 NAbs were significantly different between the two groups (0-3 weeks and 3-8 weeks) (*χ*2=14.04, *P*<0.001) (Table 2).

**Table 2.** Correlation between NAbs and the time interval between two doses of COVID-19.

| Interval between | NAb positive | NAb negative | total | *P* |
| --- | --- | --- | --- | --- |
| two doses of | n(%) | n(%) |  |  |
| vaccine |  |  |  |  |
| 0-3 Weeks | 68(63.0) | 40(37.0) | 108 |  |
| 3-8 Weeks | 773(80.8) | 207(19.2) | 980 | <0.01 |
| total | 841 | 247 | 1088 |  |

### Changes in neutralizing antibodies levels after the same person was vaccinated with COVID-19 vaccine

The neutralizing antibodies level was decreased in 31 (81.6%) and increased in 7 (18.4%) of 38 people at two time points after the second dose of vaccine. There were 6 males and 1 female in the group with elevated neutralizing antibodies level (7 patients). There were 13 males and 18 females in the group with decreased neutralizing antibodies level (31 patients). The proportion of males in the group with increased neutralizing antibody level was significantly higher than that in the group with decreased neutralizing antibody level (*χ*2=4.378, *P*<0.05). There was no significant difference in age between the group with increased neutralizing antibody level and the group with decreased neutralizing antibody level (t =-0.034, *P*>0.05).

### Linear correlation between COVID-19 NAbs and COVID-19 antibodies

Among the 1335 patients, 888 were positive for COVID-19 neutralization antibodies, with a positive rate of 66.5% (888/1335); 886 were positive for COVID-19 IgM or IgG, with a positive rate of 66.4 % (886/1335). Among the 888 patients with positive neutralization antibodies, 886 were positive for COVID-19 IgM or IgG. There was a high linear correlation between the NAbs of the new coronavirus and the antibodies of the novel coronavirus in 1335 patients (Fig 4).

**Fig 3.**
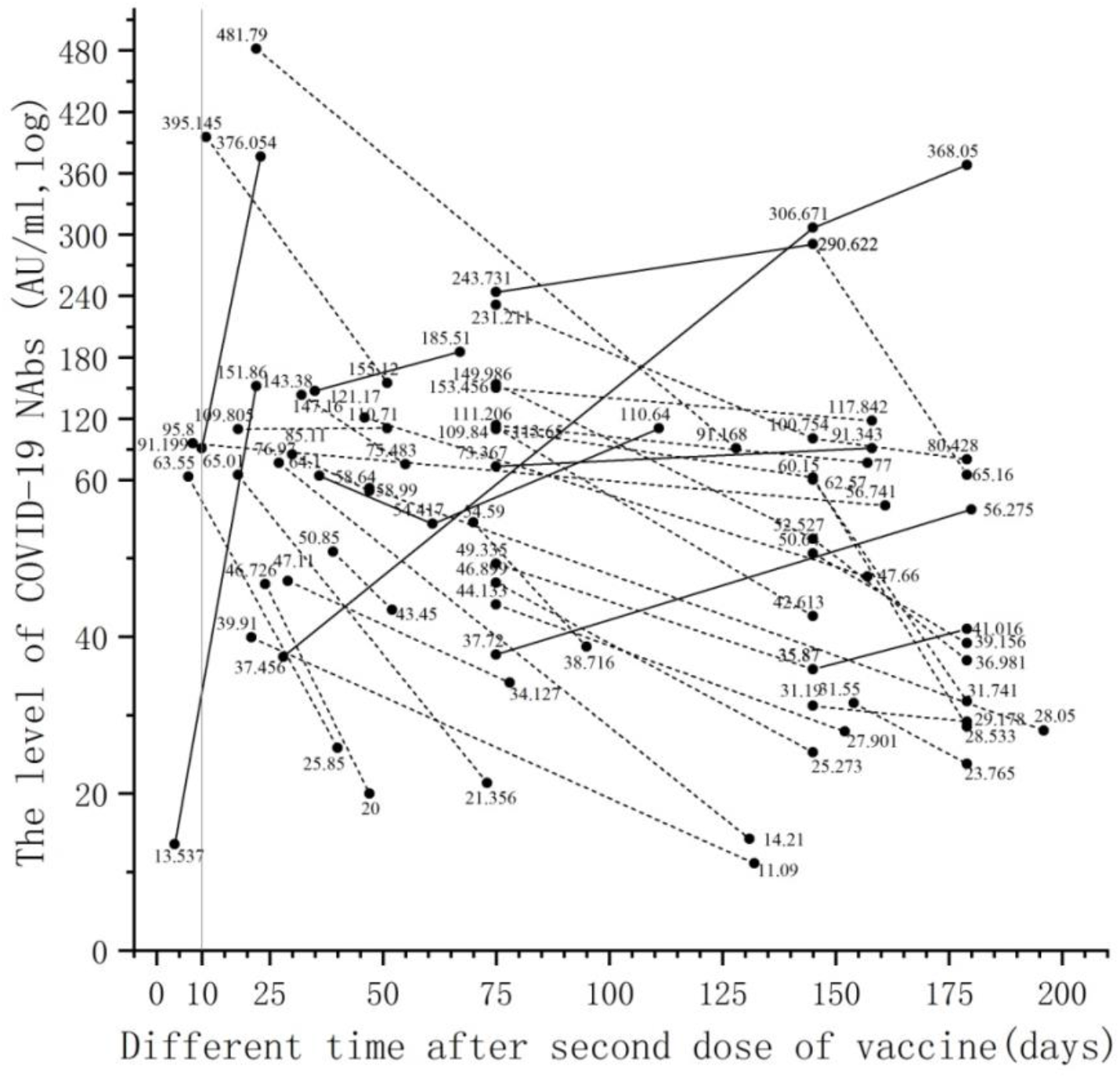
Changes in neutralizing antibodies levels between two test times after the second vaccine dose in the same person.

**Fig 4.**
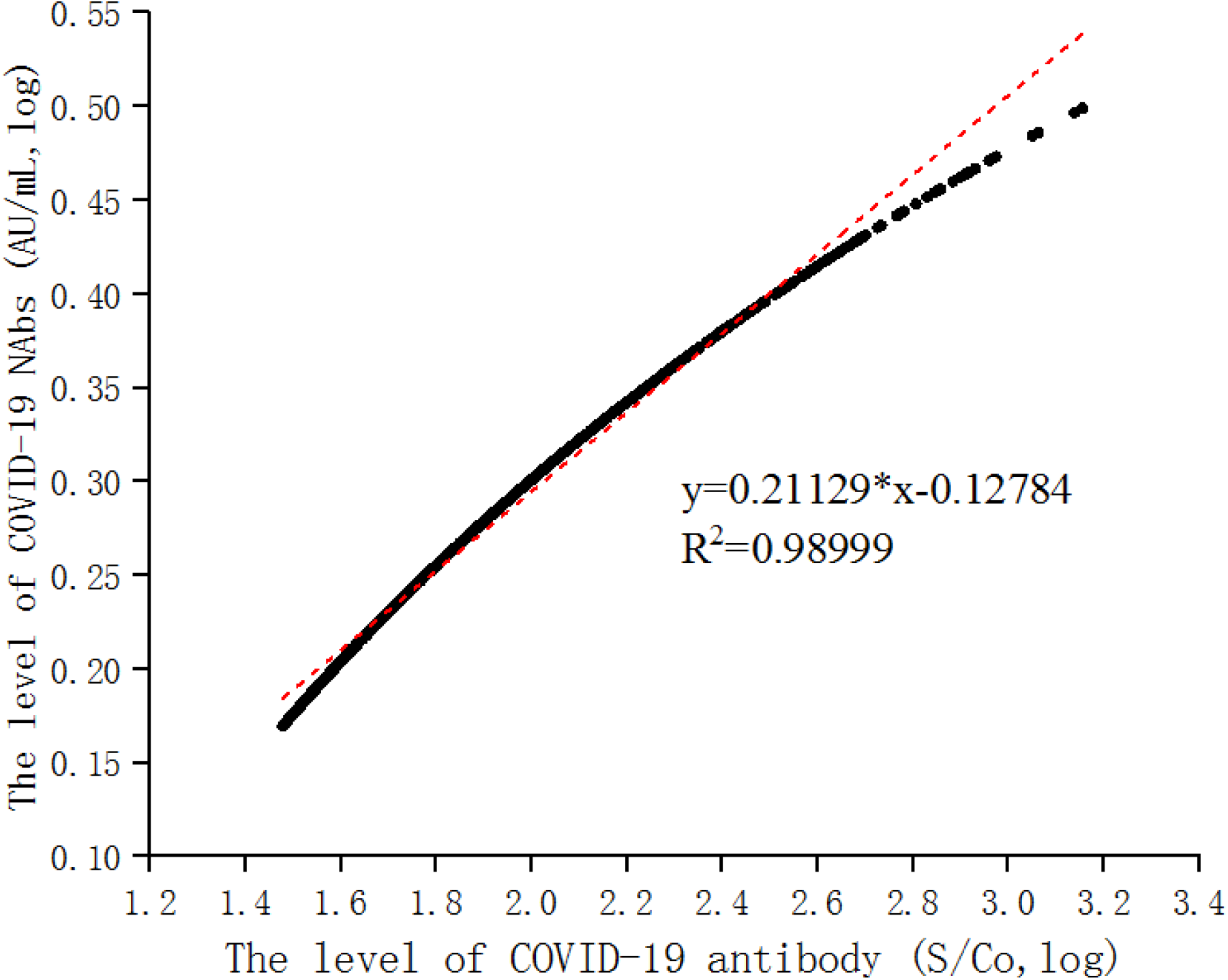
Linear correlation equation of COVID-19 neutralizing antibody and COVID-19 antibody.

**Table2.** Correlation between COVID-19 NAbs and COVID-19 antibodies.

|  | NABs positive | NABs negative | total | *P* |
| --- | --- | --- | --- | --- |
|  | N (%) | N (%) |  |  |
| COVID-19 IgM<br>or IgG positive | 886(80.1) | 219(19.9) | 1103 |  |
| COVID-19 IgM<br>or IgG negative | 2(1.7) | 228(98.3) | 232 | <0.001 |
| total | 888 | 447 | 1335 |  |

### Differences in the levels of NAbs against COVID-19 between groups receiving one dose of vaccine and two doses of vaccine

Among the 1335 patients, 1092 patients received two doses of vaccine and 243 patients received one dose of vaccine. Among 1092 patients in the two-dose vaccine group, 844 patients were positive for COVID-19 NAbs, with a positive rate of 77.3%. Among 243 cases in the one-dose vaccine group, 44 cases were positive for COVID-19 neutralizing antibody (positive rate was 18.1%); The positive rate of NAbs against COVID-19 in the two-dose vaccine group (77.3%) was significantly higher than that in the one-dose vaccine group (18.1%), with statistical difference (*χ*2=312.590, P<0.001) (Table 3).

**Table 3.**
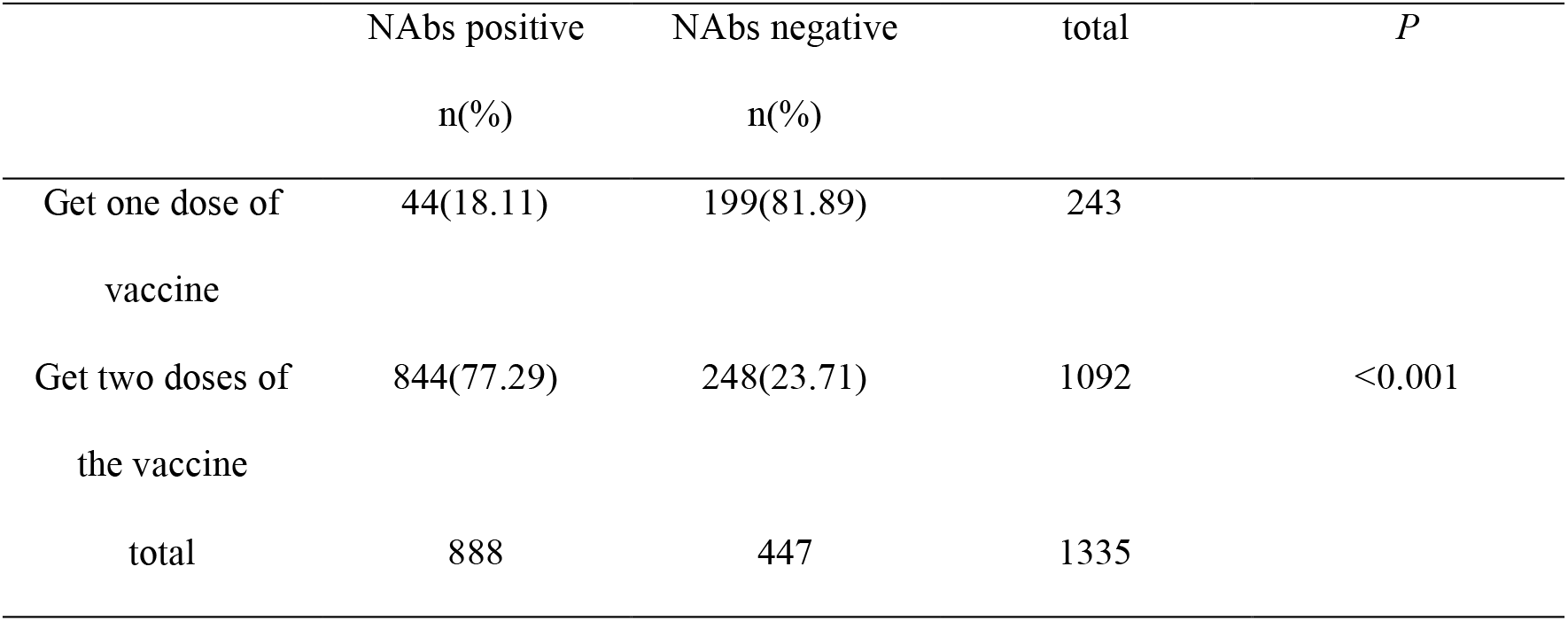
Correlation between NAbs and number of vaccinations.

|  | NAbs positive<br>n(%) | NAbs negative<br>n(%) | total | *P* |
| --- | --- | --- | --- | --- |
| Get one dose of vaccine | 44(18.11) | 199(81.89) | 243 |  |
| Get two doses of the vaccine | 844(77.29) | 248(23.71) | 1092 | <0.001 |
| total | 888 | 447 | 1335 |  |

### Correlation between age and gender and the level of NAbs against COVID-19 in the two-dose vaccine group

A total of 206 people who were 11-70 days after receiving the second dose were tested and divided into three groups: 18-40 years old, 41-60 years old and >60 years old. The positive rates of NAbs in the three groups were 95.14%, 78.43% and 81.8%, respectively. The positive rate of neutralizing antibody was significantly higher in 18 to 40 years old than in 41 to 60 years old (*χ*2=12.547, *P* <0.01). There was no significant difference in the positive rate of NAbs between 18 to 40 years old and >60 years old, and between 41 to 60 years old and >60 years old (*χ*2=3.315, *P* >0.01; *χ*2=0.063, *P* >0.01). The titer of neutralizing antibody in 18-40 years old group was significantly higher than that in 41-60 years old group (t=-0.222, *P* <0.01). The titer of NAbs was slightly lower in the 41-60 years old group than in the >60 years old group, but there was no significant difference (t =-0.010, *P* >0.05). The titer of NAbs produced in 18-40 years old group was lower than that in >60 years old group, with no significant difference (t=-1.41, *P* >0.05) (Table 4).There was a poor linear correlation between age and antibody titers in 1335 patients(Fig 5).

**Table 4.** Differences in the levels of COVID-19 NAbs in different age groups vaccinated with the two inactivated vaccines.

|  | NAbs positive | NAbs negative | total |
| --- | --- | --- | --- |
|  | n(%) | n(%) |  |
| 18-40y | 137(95.14) | 7(4.86) | 144 |
| 41-60y | 40(78.43) | 11(21.57) | 51 |
| >60y | 9(81.82) | 2(18.18) | 11 |
| total | 186 | 20 | 206 |

**Fig 5.**
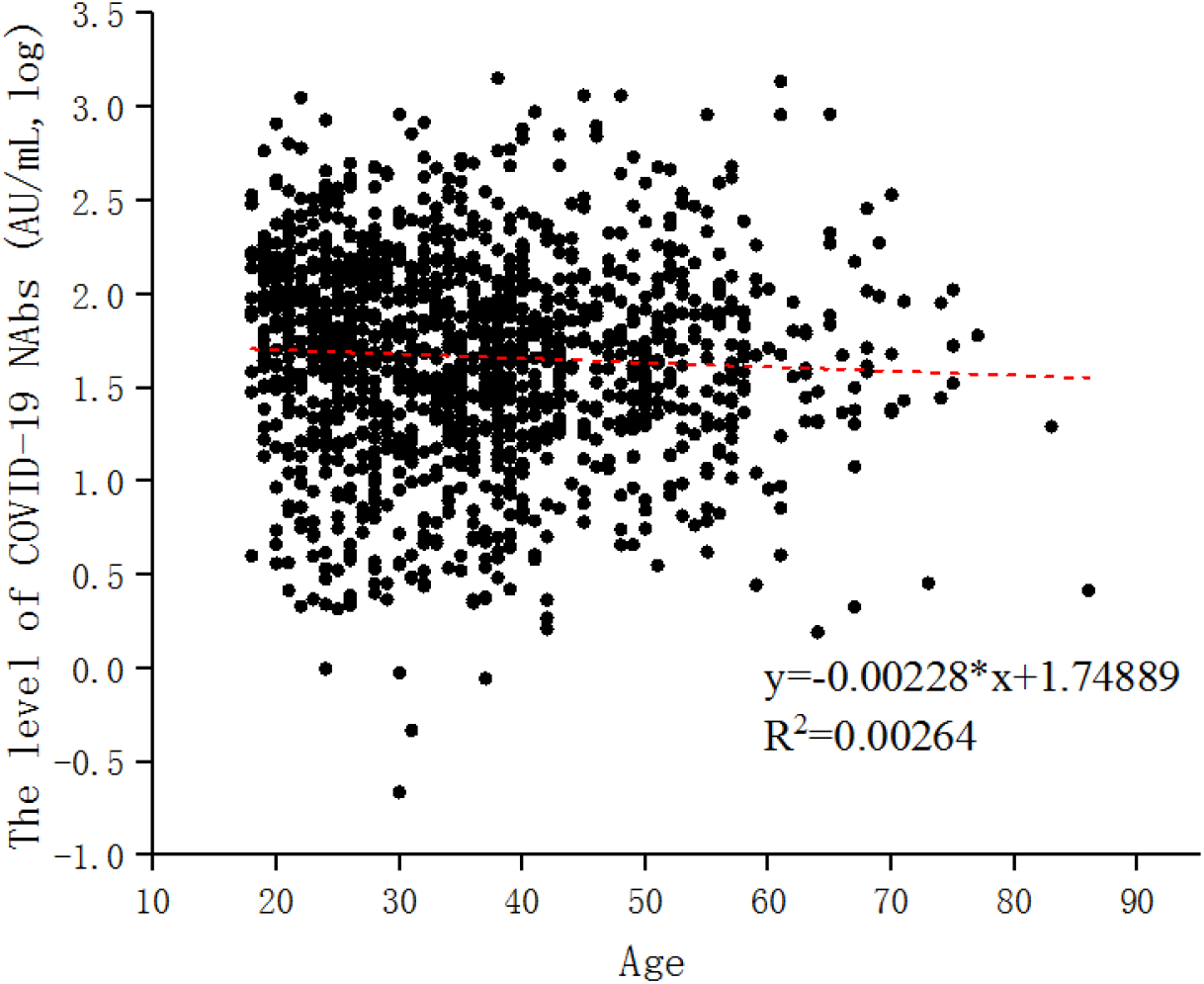
The linear correlation equation between age and COVID-19 neutralizing antibody.

A total of 206 people who were 11-70 days after the second injection were detected and divided into male and female groups. The positive rate of NAbs in male group (89.32%) was lower than that in female group (91.26%), but there was no significant difference (*χ*2=0.222, *P* >0.05). The titer of NAbs in male group was higher than that in female group, but there was no significant difference (t =0.081, *P* >0.05) (Table 5).

**Table 5.** Differences in the levels of COVID-19 NAbs in different age groups vaccinated with the two inactivated vaccines.

|  | NAbs positive | NAbs negative | total | *P* |
| --- | --- | --- | --- | --- |
|  | N (%) | N (%) |  |  |
| Male | 92(89.32) | 11(10.68) | 103 |  |
| Female | 94(91.26) | 9(8.74) | 103 | >0.05 |
| Total | 186 | 20 | 206 |  |

## Discussion

Our study found that the positive rate of NAbs was 66.5% in adults who received one or two doses of inactivated vaccine and 77.3% in adults who received two doses of inactivated vaccine. In a phase 3 randomized clinical trial of adults vaccinated with Chinese inactivated vaccines, the protective efficacy of two whole virus inactivated vaccines was 72.8% and 78.1%, respectively [5]. Our study two dose inactivated vaccine inoculation adult produce NAbs positive rate is similar to previous studies, but one or two doses inactivated adult produce NAbs positive rate was 66.5%, slightly lower than that of previous studies, may because of our research, 18.1% of vaccinated with only one dose, and previous research is two vaccine inoculation. The phase 3 clinical trials reported in the literature mainly included healthy and young people from the Middle East and other Asian countries, and the Middle East was limited to the male population. Our study reflects the level of NAbs induced by vaccination in the real world of our population. Recently published interim results from phase 3 clinical trials of other vaccines, these included two mRNA vaccines (BNT162b2 and mrN-1273) and three adenovirus-based vaccines (ChAdOx1 nCoV-19, GAM-COVID-19 [Sputnik V] and Ad26.COV2.S) [4,8-11]. The protective efficacy of BNT162b2 (Pfizer-BionTech) vaccine was reported to be 95.0%; The protective efficacy of mrNA-1273 (Moderna) vaccine was 94.1%; ChAdOx1 nCoV-19 62.1% and 90.0%, depending on dose regimen (2 standard doses or low doses followed by standard doses); For GAM-COVID-19 vaccine, 91.6%; The AD26.COV2.S vaccine was 66.9%. These studies mainly include people from western countries, the proportion of the aged and participants is higher, our study is the Chinese people, ethnic and regional different may lead to a vaccine induced neutralizing antibody level is different, therefore, our research can response more Chinese people to produce the actual situation of neutralizing antibody after vaccination.

The WHO international standard for SARS-CoV-2 NAbs was developed by the National Institute for Biological Standards and Control (NIBSC) in collaboration with the WHO and is the final reference substance for national standards. The WHO released the first-generation international standard (NIBSC Code: 20/136) for COVID-19 antibodies through collaborative calibration among international laboratories, and determined the international unit/mL (IU/mL) concentration of NAbs, laying a foundation for subsequent quantitative studies. International standard (IS) considered to be the highest level of biological material reference materials, the international standard for IU quantitative sample for the unit of biological activity, which can be compared from different laboratory analysis and make the results comparable to better define the analysis parameters (e.g., sensitivity of the test) and clinical parameters (such as the protection of the antibody level). SARS-CoV-2 Neutralizing antibody IS will facilitate the standardization of serological assays for detection of SARS-CoV-2 for use in vaccinology studies to detect antibodies produced by human vaccination. Compared with the original WHO value, the coefficient of variation (CV) of our study was 1.3%, which met the WHO standard of less than 3%. Therefore, our neutralizing antibody unit AU/mL is close to the IU/mL and will help to evaluate vaccine efficacy and data from epidemiological and immunological surveillance studies.

An important question in the efficacy and safety of COVID-19 vaccines is how long the protection they provide will last. Our results show that the positive rate of NAbs is 82%-100% on days 11-70, after which the positive rate starts to decline. Therefore, it is recommended to test the neutralizing antibody level from day 10-70 after the second dose of vaccine. on days 71-332, the positive rate of COVID-19 NAbs drops to 27%, so in the real world, the level of COVID-19 NAbs drops from days 61-70 and a third dose of vaccine is recommended. The New England Journal of Medicine published persistent data on the immune response stimulated by the candidate COVID-19 vaccine Moderna mRNA-1273 [12]. Analysis showed that participants maintained high levels of antibodies binding to the Novel Coronavirus spike protein and NAbs 90 days after the second dose of vaccine. Even if the level of NAbs decreases slightly over time, mRNA-1273 still has the potential to provide lasting humoral immunity. In addition, a study published in Science [13] showed that even with low plasma neutralizing antibodies activity triggered by the Novel Coronavirus natural infection, it could still trigger a robust memory B cell response. When encountering Novel Coronavirus again, these memory B cells can quickly produce targeted NAbs, which is crucial for maintaining long-term immunity to Novel coronavirus. Different studies on the levels of NAbs in rehabilitated patients show that the persistence of NAbs in different patients is uneven. In some patients, the level of NAbs decreased significantly within a few months after recovery, so we studied the persistence of neutralizing antibody levels triggered by COVID-19 vaccine.

Wang Huaqing, chief expert of the Immunization program of the Chinese Center for Disease Control and Prevention, said at a press conference under the Joint prevention and control mechanism of The State Council that the interval between inactivated vaccine injections should be completed within eight weeks[14]. In the two-dose vaccine group, the time interval was from 0 days to 77 days, and the highest level of NAbs generated at the interval of 21-56 days between the two doses suggested that 21-56 days between the two doses was suitable for vaccination.

People aged 60 and above are the people with severe illness and high risk of death after contracting novel coronavirus. Data from phase I/II clinical studies showed that novel coronavirus vaccine was safe in this population[15], and our study found that the average production of NAbs in the elderly (>60 years old) was lower than that in the young (18-40 years old), which was also confirmed by previous studies [15]. Instead of receiving inactivated vaccine in this study, the study population was vaccinated with mRNA vaccine. Our study also found that the positive rate of NAbs in men in the two-dose group was significantly lower than in women, but the difference was not significant. Our study suggested the level of NAbs produced after vaccination was affected by age, but not by gender.

The gold standard method for NAbs is the neutralization test[16]. Even if live or synthetic viruses are used to react with samples to detect the killing ability of NAbs against viruses in samples, the clinical efficacy evaluation of vaccines published now is all using this method [5, 8]. However, this method has high requirements for testing and is not suitable for general medical testing institutions. Another kind of NAbs detection is to use the immune reaction principle, through the specific antigen detection corresponding neutralizing antibody, this method is suitable for general medical testing institutions to promote the use. Because now the epidemic strains are just some point mutations on individual sites, there are mainly five or six mutations, and there are many neutralizing antibody on the S protein epitope, individual point mutations will not lead to virus escape completely, that’s why we also effective vaccine, the vaccine on RBD area S protein is the main work of neutralizing antibody table, Our neutralizing antibody reagent uses the S protein RBD, which is detectable in the same way that vaccines work. Our study showed a high linear correlation between NAbs and COVID-19 IgM/IgG antibodies in vaccinated individuals.

Therefore, it is suggested that when NAbs cannot be detected, COVID-19 IgM/IgG antibodies can be detected instead. 89.6% (796/888) of these IgG positive samples were also positive in neutralisation tests(NT) in our study. This result differs from the results of a recently published study by Müller K et. al., in which only 68.7% (158/230) of these IgG positive samples were also positive in NTs [17]. This paper uses ELISA method to detect COVID-19 antibody, which is different from chemiluminescence method used in our paper. Antibodies against SARS-CoV-2 produced by convalescent patients reduced the neutralization effectiveness of emerging mutations. So it’s critical that we monitor SARS-CoV-2 antibody levels for vaccine surveillance and vaccine development[18].

Limitations of this study. For one thing, the study did not include pregnant women or people under 18 years old. Therefore, the real-world effectiveness of inactivated vaccines in these populations remains unknown. In the next step, the effective protective concentration of COVID-19 neutralizing antibody can be determined through research, for example, the critical value of hepatitis B virus surface antibody is 10 mIU/mL.

In summary, the positive rate of neutralization antibody was the highest at 10-70 days after the second dose of vaccine, and gradually decreased with the passage of time. Therefore, the third dose of vaccine was recommended at about 70 days.our study shows that COVID-19 IgM/IgG antibodies can be detected in cases where NAbs cannot be detected. 21-56 days(3-8 week) between the two doses was suitable for vaccination. Our study provides an important insight into real-world vaccination cycles, vaccination timing, and detection methods.

## Data Availability

All data used to draw the conclusions in this paper are provided in the paper and/or in the supplementary material.

## References

1. Draft landscape and tracker of COVID-19 candidate vaccines. World Health Organization. Accessed May 18, 2021. https://www.who.int/publications/m/item/draft-landscape-of-COVID-19-candidate-vaccines.

2. Kyriakidis NC, López-Cortés A, González EV,Grimaldos AB, Prado EO. SARS-CoV-2 vaccines strategies: a comprehensive review of phase 3 candidates. NPJ Vaccines. 2021;6(1):28.

3. Xia S, Duan K, Zhang Y, Zhao DY, Zhang HJ, Xie ZQ, et al. Effect of an inactivated vaccine against SARS-CoV-2 on safety and immunogenicity outcomes: interim analysis of 2 randomized clinical trials. JAMA. 2020;324(10):951–960.

4. Baden LR, El Sahly HM, Essink B, Kotloff K, Frey S, Novak R, et al. Efficacy and safety of the mRNA-1273SARS-CoV-2 vaccine. N Engl J Med. 2021;384(5):403–416.

5. Al Kaabi N, Zhang Y, Xia S, Yang Y, Al Qahtani MM, Abdulrazzaq N, et al. Effect of 2 Inactivated SARS-CoV-2 Vaccines on Symptomatic COVID-19 Infection in Adults: A Randomized Clinical Trial. JAMA. 2021, 326(1):35–45.

6. http://www.nhc.gov.cn/xcs/yqfkdt/202108/39679e47dbb849e386f64247fd241082.shtml

7. Long QX, Tang XJ, Shi QL, Li Q, Deng HJ, Yuan J, et al. Clinical and immunological assessment of asymptomatic SARS-CoV-2 infections. Nature Medicine. 2020; 26(8):1200–1204.

8. Polack FP, Thomas SJ, Kitchin N, Absalon J, Gurtman A, Lockhart S, et al; Safety and efficacy of the BNT162b2 mRNA COVID-19 vaccine. N Engl J Med. 2020;383(27):2603–2615.

9. Logunov DY, Dolzhikova IV, Shcheblyakov DV, Tukhvatulin AI, Zubkova OV, Dzharullaeva AS, et al; Safety and efficacy of an rAd26 and rAd5 vector-based heterologous prime-boost COVID-19 vaccine: an interim analysis of a randomised controlled phase 3 trial in Russia. Lancet. 2021;397(10275):671–681.

10. Voysey M, Clemens SAC, Madhi SA, Weckx LY, Folegatti PM, Aley PK, et al; Safety and efficacy of the ChAdOx1 nCoV-19 vaccine (AZD1222) against SARS-CoV-2: an interim analysis of four randomised controlled trials in Brazil, South Africa, and the UK. Lancet. 2021; 397(10269):99–111.

11. Sadoff J, Gray G, Vandebosch A, Cárdenas V, Shukarev G, Grinsztejn B, et al; Safety and efficacy of single-dose Ad26.COV2.S vaccine against Covid-19. N Engl J Med. 2021, 384(23):2187–2201.

12. Widge AT, Rouphael NG, Jackson LA, Anderson EJ, Roberts PC, Makhene M, et al. Durability of Responses after SARS-CoV-2 mRNA-1273 Vaccination. N Engl J Med. 2021, 384(1):80–82.

13. Dan JM, Mateus J, Kato Y, Hastie KM, Yu ED, Faliti CE, et al. Immunological memory to SARS-CoV-2 assessed for up to 8 months after infection. Science. 2021,371(6529): eabf4063.

14. https://baijiahao.baidu.com/s?id=1700327464370422756&wfr=spider&for=pc.

15. Walsh EE, Frenck RW Jr, Falsey AR, Kitchin N, Absalon J, Gurtman A, et al. Safety and Immunogenicity of Two RNA-Based Covid-19 Vaccine Candidates. N Engl J Med. 2020. 383(25): 2439–2450.

16. María Elvira Balcells, Luis Rojas, Nicole Le Corre, Constanza Martínez-Valdebenito, María Elena Ceballos, Marcela Ferrés, et al. Early versus deferred anti-SARS-CoV-2 convalescent plasma in patients admitted for COVID-19: A randomized phase II clinical trial. PLoS Med. 2021, 18(3): e1003415.

17. Müller K, Girl P, von Buttlar H, Dobler G, Wölfel R. Comparison of two commercial surrogate ELISAs to detect a neutralising antibody response to SARS-CoV-2. J Virol Methods. 2021;292:114122.

18. Fiona Tea, Alberto Ospina Stella, Anupriya Aggarwal, David Ross Darley, Deepti Pilli, Daniele Vitale, et al. SARS-CoV-2 neutralizing antibodies: Longevity, breadth, and evasion by emerging viral variants. PLoS Med. 2021, 18(7): e1003656.

